# The Impact of Fontan Circulatory Failure on Heart Transplant Survival: A 20 Center Retrospective Cohort Study

**DOI:** 10.1101/2024.11.13.24317295

**Authors:** Kurt R Schumacher, David N Rosenthal, Adriana Batazzi, Sunkyung Yu, Garrett Reichle, Maria Bano, Shriprasad R Deshpande, Matthew O’Connor, Humera Ahmed, Sharon Chen, Lydia K. Wright, Steven J Kindel, Anna Joong, Michelle Ploutz, Brian Feingold, Justin Godown, Chad Y Mao, Angela Lorts, Kathleen E. Simpson, Aecha Ybarra, Marc E Richmond, Shahnawaz Amdani, Jennifer Conway, Elizabeth D Blume, Melissa K Cousino

## Abstract

**Background:** Fontan circulatory failure (FCF) is a chronic state in palliated single ventricle heart disease with high morbidity and mortality including heart failure, multisystem end-organ disease, and need for heart transplant. Specific FCF morbidities have not been rigorously defined, limiting study of how FCF morbidities impact pre- and post-HT outcomes. We hypothesized that FCF-related morbidities affect survival from heart transplant waitlisting through 1-year post-heart transplant.

**Methods:** This 20-center, retrospective cohort study collected demographic, medical/surgical history, waitlist data, and peri- and post-heart transplant data, and *a priori* defined FCF-specific morbidities in Fontan patients who were listed for heart transplant from 2008-2022. Univariate 2-group statistics compared surviving individuals with those who 1) died anytime from waitlisting to 1-year post-heart transplant, 2) died on the waitlist, 3) underwent transplant and died within 1-year post-transplant. Using covariates from both univariate analyses, multivariable logistic regression determined the primary study outcome of independent FCF risk factors for mortality between waitlist and 1-year post-heart transplant

**Results:** Of 409 waitlisted patients, 24 (5.9%) died on the waitlist. Of the 341 (83.4%) who underwent HT, 27 (8.5%) did not survive to 1-year. Univariate risk factors for waitlist death included higher aortopulmonary collateral burden, > 1 hospitalization in prior year, younger age, sleep apnea, higher NYHA class, non-enrollment in school or work, and single-parent home. Risk factors for 1-year post-heart transplant mortality included hypoplastic left heart syndrome diagnosis, patent fenestration, anatomic Fontan obstruction, clinical cyanosis (pulse oximetry < 90%), polycythemia, portal variceal disease, mental health condition requiring treatment, and higher HLA class II PRA. Of the patients not surviving from waitlisting to 1-year post-heart transplant, independent risk factors for mortality included >1 hospitalization in the year prior to waitlisting (adjusted odds ratio 2.0, p=0.05) and clinical cyanosis (adjusted odds ratio 5.0, p=0.002).

**Conclusions:** Patients with Fontan palliation selected for heart transplant have significant mortality from waitlisting through transplant. Among FCF specific morbidities, cyanosis is associated with worsened survival and necessitates further study. Clinical morbidity of any type requiring repeated hospital admission also should prompt consideration of heart transplant.

**Clinical Perspective:** *What is new?:*

- Survival through heart transplant in patients with Fontan physiology selected for waitlisting has increased from previous reports, but this patient group still has significant risk of mortality.
- Risk factors for waitlist mortality and post-transplant mortality are different.
- Cyanosis and repeated hospitalizations prior to listing are independent risk factors for mortality between waitlisting and 1-year post-heart transplant.

*What are the clinical implications?:*

- To successfully manage a patient through the entire transplant process, attention to mitigating different risks in the waitlist and post-transplant phase is necessary.
- Repeated hospitalization or significant cyanosis in a patient with Fontan physiology should prompt consideration of heart transplant.

## Introduction

Fontan circulatory failure (FCF) is a multisystem syndrome directly resulting from the physiologic ramifications of staged surgical palliation to total cavopulmonary anastomosis, or the Fontan palliation, for single ventricle type congenital heart diseases (CHD). Fontan palliation offers individuals born with single ventricle CHD long-term survival with the alleviation of cyanosis, but it also results in lifelong venous hypertension which may be accompanied by compromise in cardiac output and other cardiovascular abnormalities. Since every organ is exposed to this abnormal circulation, unique and Fontan-specific multisystem abnormalities have been described in the recent era. These abnormalities do not occur in isolation, making FCF effectively a systemic disease that is associated with poor long-term survival [1, 2].

In an individual with Fontan circulation, heart transplant removes Fontan physiology and restores normal biventricular circulation, thus it is used as a definitive therapy for FCF. Unfortunately, individuals with FCF have reported post-heart transplant mortality that exceeds that of most other types of heart disease [3, 4]. This is likely impacted by the residual systemic implications of Fontan physiology as well as uncertainty surrounding optimal timing for heart transplant evaluation referral in this lifelong chronic disease state [5]. The differential survival between FCF and non-FCF patients is seen in the early post-heart transplant period. For patients who survive the first-year, outcomes are similar or superior to those with other heart diseases undergoing heart transplant ([4].

The risk of mortality being most pronounced perioperatively and early post-HT suggests that pre-operative FCF characteristics influence outcome. Attempts to study the FCF- transplant interaction have been limited by inadequate sample size or datasets with marked lack of FCF-specific, granular data. A recent multicenter study in adults with Fontan suggest FCF states may be important but did not fully characterize the unique,

Fontan-specific multisystem effects of FCF [6]. As a result of these limitations, it remains unclear which FCF disease states affect outcomes.

This retrospective study aimed to fully characterize patients with FCF who were waitlisted for heart transplant by applying FCF-specific morbidity definitions, which were developed via Delphi methodology and previously described [7], to a large, multicenter sample. It was hypothesized that FCF-specific morbidities would affect survival from heart transplant waitlisting through 1-year post-heart transplant.

## Methods

### Study Design and Population

This is a retrospective cohort study of patients with Fontan-palliated single ventricle CHD who underwent waitlisting for heart transplant between 2008 and 2022 at twenty congenital heart transplant centers in the United States and Canada. Both pediatric patients and adults were eligible for inclusion. Multi-organ transplant patients were also eligible for inclusion. Patients who were evaluated for transplant but never active on the waitlist were not included as retrospective center capture of this population was variable and incomplete. The study was approved by the Institutional Review Board at the primary coordinating center at the University of Michigan as well as the Institutional Review Boards of all 20 participating centers.

### Data Extraction

De-identified patient-level data were extracted through medical chart review and entered in HIPPA-compliant REDCap. Demographic data included sex, race, age, congenital heart disease diagnosis, and surgical history. At time of waitlisting, extracted data included NYHA class, history of hospitalizations in the year prior to waitlisting, multiple serum laboratories, and clinical testing data (e.g., echocardiogram, cardiac MRI, cardiac catheterization, cardiopulmonary exercise testing, 6-minute walk testing results). Data were also extracted on events that occurred during the waitlisting period including initiation of new vasoactive treatments, invasive ventilation, new ventricular assist device placement, support with extracorporeal membrane oxygenation, and waitlist actions including removal from the waitlist and reason for removal. Mortality at any point following listing and major post-transplant morbidities including rejection and infection in the first post-transplant year were also collected.

### Defining FCF-Specific Morbidities

Previously described FCF morbidity definitions developed via modified Delphi methodology were applied to the extracted dataset [7]. Although the published definitions included graded severities for each morbidity, each morbidity was treated dichotomously as ‘present’ or ‘absent’ at the time of waitlisting for the purpose of this initial study due to the high number of variables in addition to FCF morbidities already being considered in the analysis.

### Outcomes

The primary outcome was death at any time between waitlisting and 1-year post-heart transplant. Secondary outcomes included death prior to heart transplant (which included patients removed from the waitlist due to deterioration in clinical status) and death in a transplanted patient within 1-year post-transplant.

### Statistical Analysis

Data are reported as frequency and percentage (%) for categorical variables and median with interquartile range (IQR) or mean ± standard deviation (SD) for continuous variables. Univariate associations of patient and clinical characteristics including a *priori* defined FCF-specific morbidities with each of the study outcomes (waitlist death, death within 1-year post-transplant, and death at any time between waitlisting and 1-year post-heart transplant) were examined using Chi-square test or Fisher’s exact for categorical variables and Wilcoxon rank-sum test or two-sample t-test for continuous variables. Due to the limited number of cases of either waitlist mortality or 1-year post-transplant mortality, multivariable analyses for those outcomes were not feasible. Specifically, because of the study’s *a priori* intent to define risk of both post-waitlist and post-transplant death as a single, unique outcome, multivariable logistic regression was used to assess independent risk factors associated with mortality between waitlisting and 1- year post-transplant after excluding individuals still on the waitlist or de-listed from the analysis. Factors found to be associated with mortality between waitlisting and 1-year post-transplant in univariate analysis (p < 0.15) were further considered and included in multivariable analyses. Due to the limited number of outcomes, the final model was compiled after a series of prior models, each analyzing a limited number of covariates, were explored to aid in variable selection and avoid overfitting. Multicollinearity among candidate variables for multivariable analysis was examined using Variance Inflation Factor. Adjusted odds ratios (aORs) with 95% confidence intervals from multivariable logistic regression were reported. Kaplan-Meier curves were used to estimate survival from waitlist to transplant, survival in the first-year post-transplant, and overall survival since waitlisting. All analyses were performed using SAS Version 9.4 (SAS Institute Inc., Cary, NC), with statistical significance set at a p-value < 0.05 using a two-sided test.

Authors KRS and MKC had full access to all data in the study and take responsibility for its integrity and the data analysis.

## Results

In total, 409 individuals with Fontan listed for heart transplant were included in the study cohort. The cohort’s demographic and prior surgical characteristics are presented in Table 1 as well as a consort diagram of the cohort (Figure 1)

**Figure 1.**
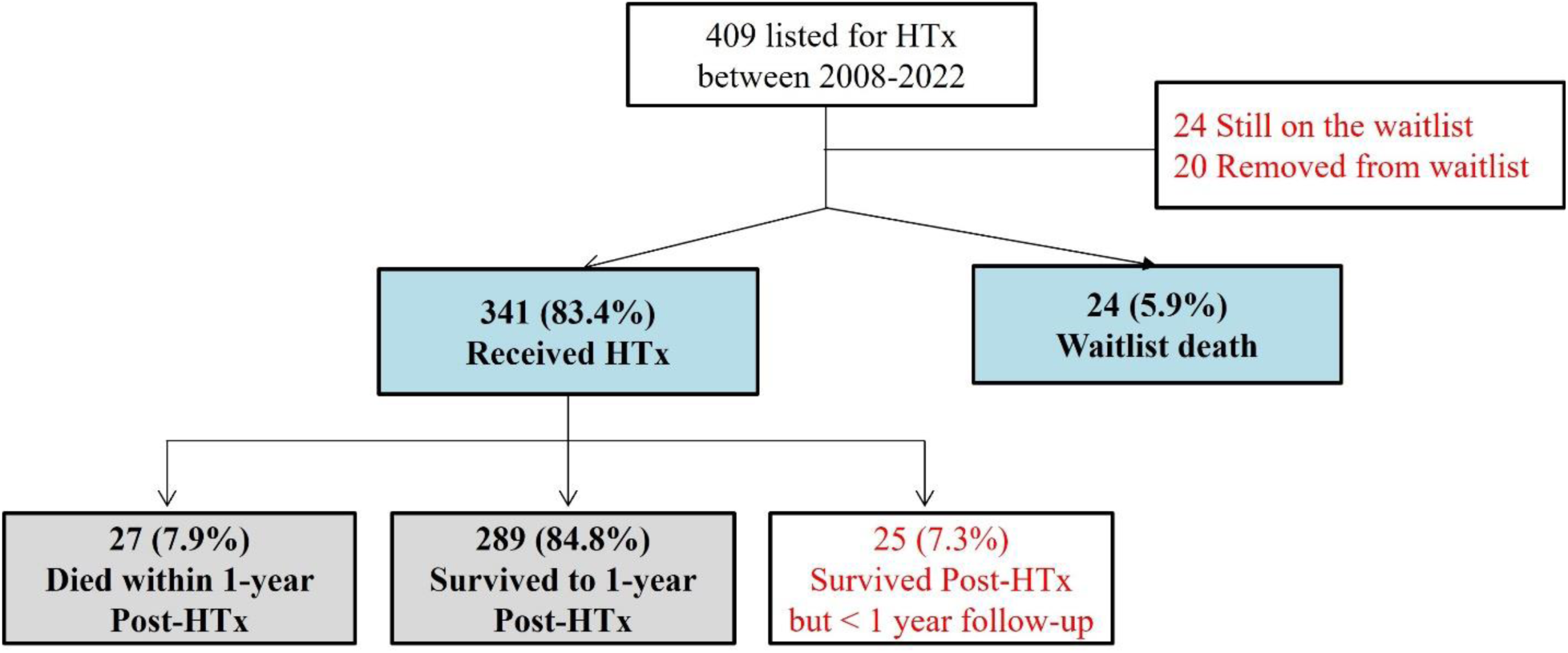
CONSORT diagram for the cohort.

**Table 1.**
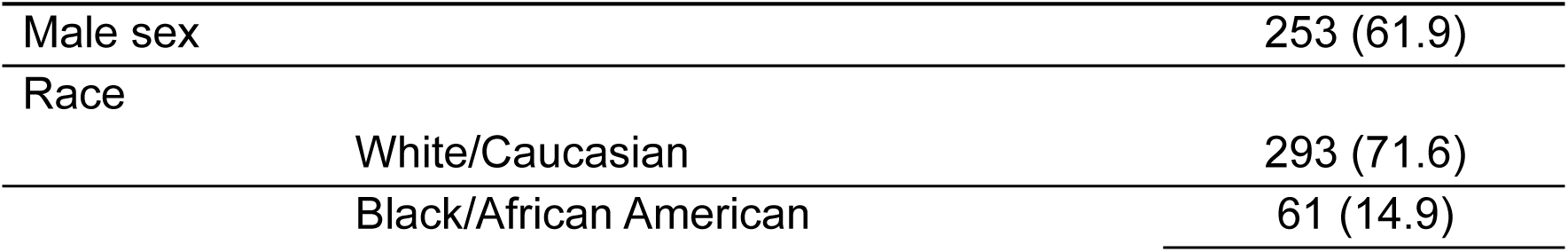
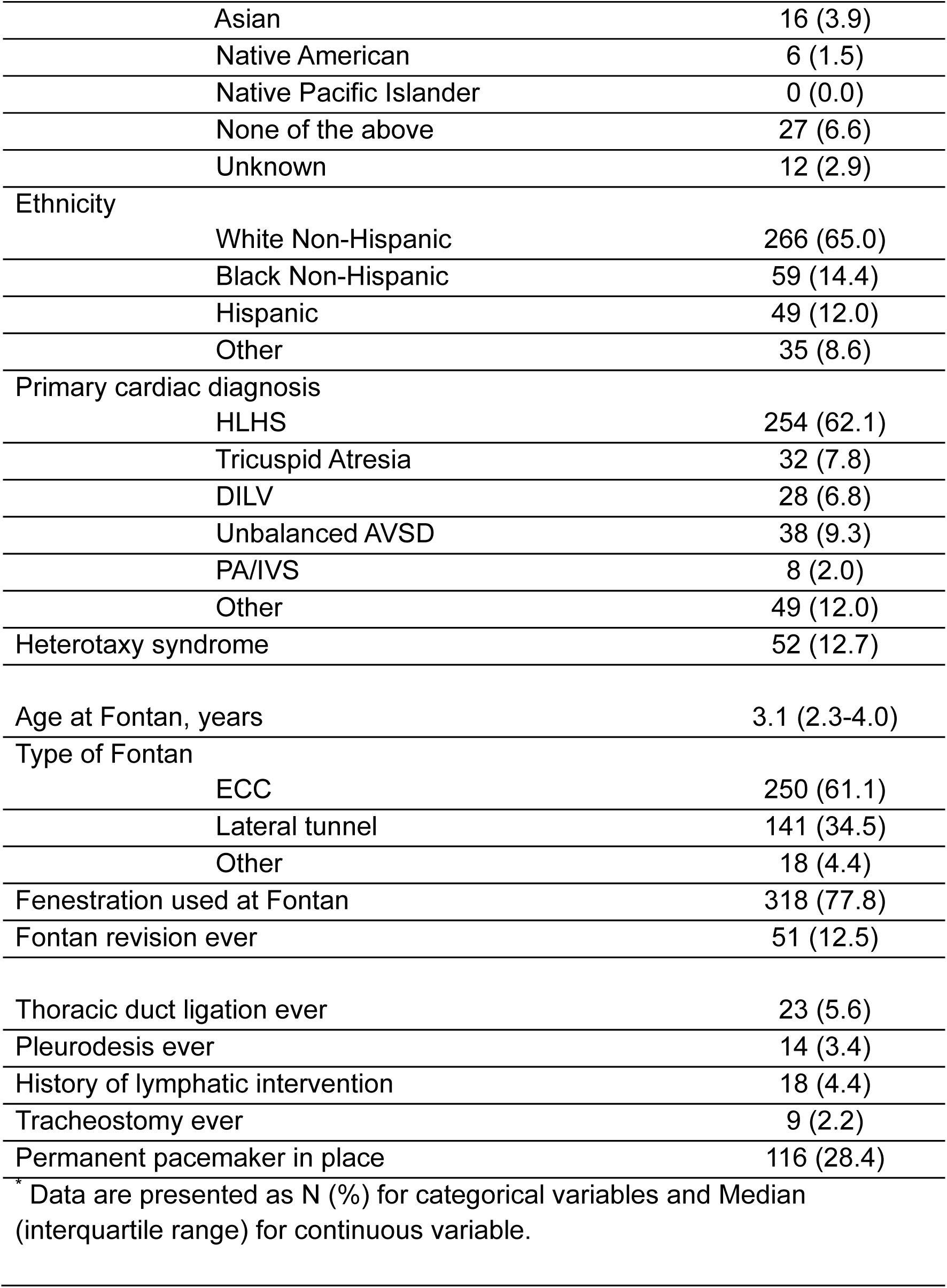
Demographics and Surgical information (N=409)

### Fontan patient characteristics at waitlisting and waitlist outcomes

The median total follow-up time post-waitlisting for the cohort was 4.2 years (IQR 2-7.3). Individual and testing characteristics at waitlisting are shown in Table 2. Additionally, cPRA data was available for 310 (75.8%) patients of the cohort; the majority did not exhibit serum anti-HLA antibodies but there was significant variation. The median total cPRA was 0% with IQR of 0-22% and a total range of 0-100%. The median class I cPRA was 0%, IQR 0-14%, total range 0-100%, and the median class II cPRA was 0%, IQR 0-10%, range 0-100%.

**Table 2.**
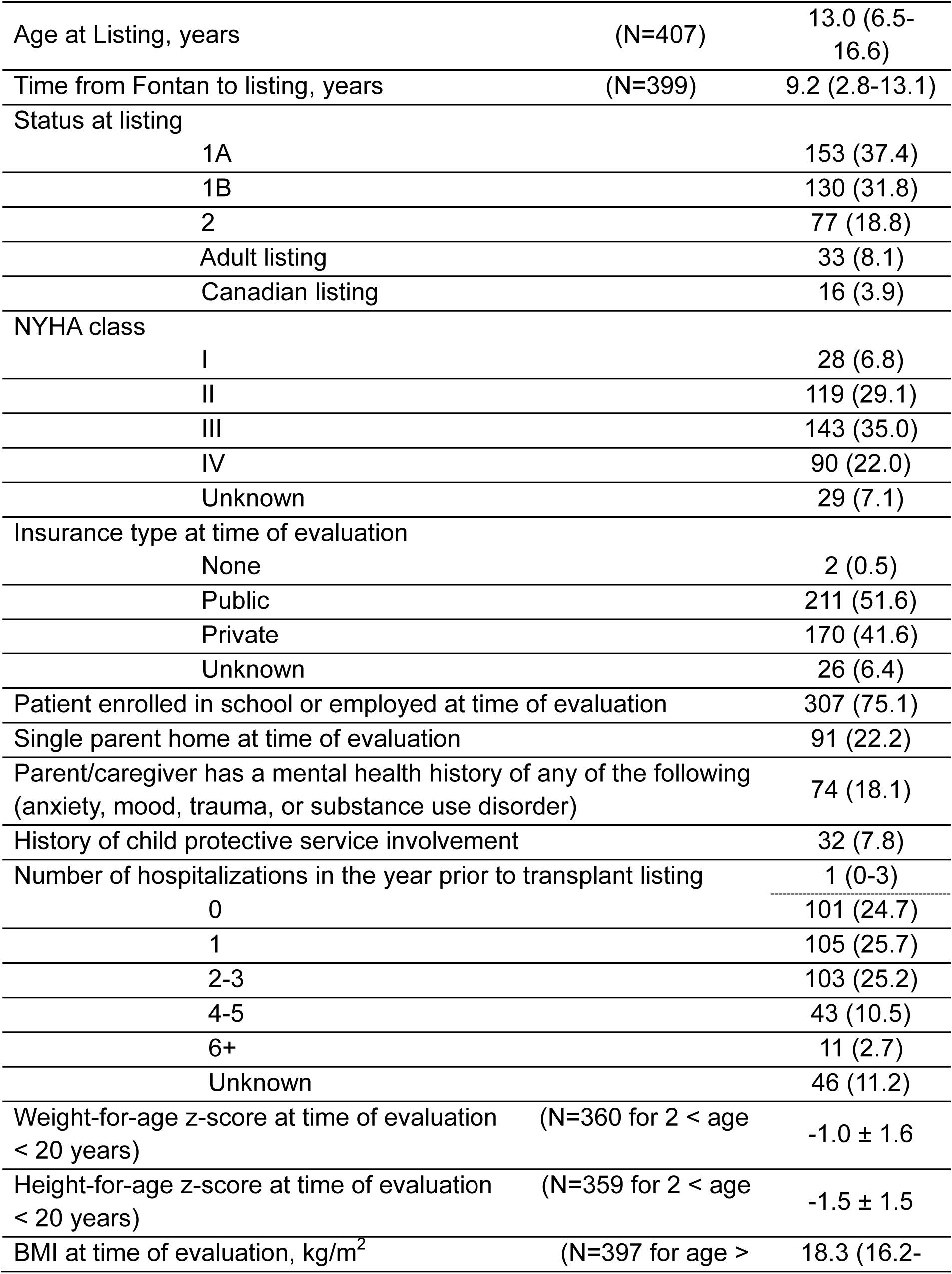
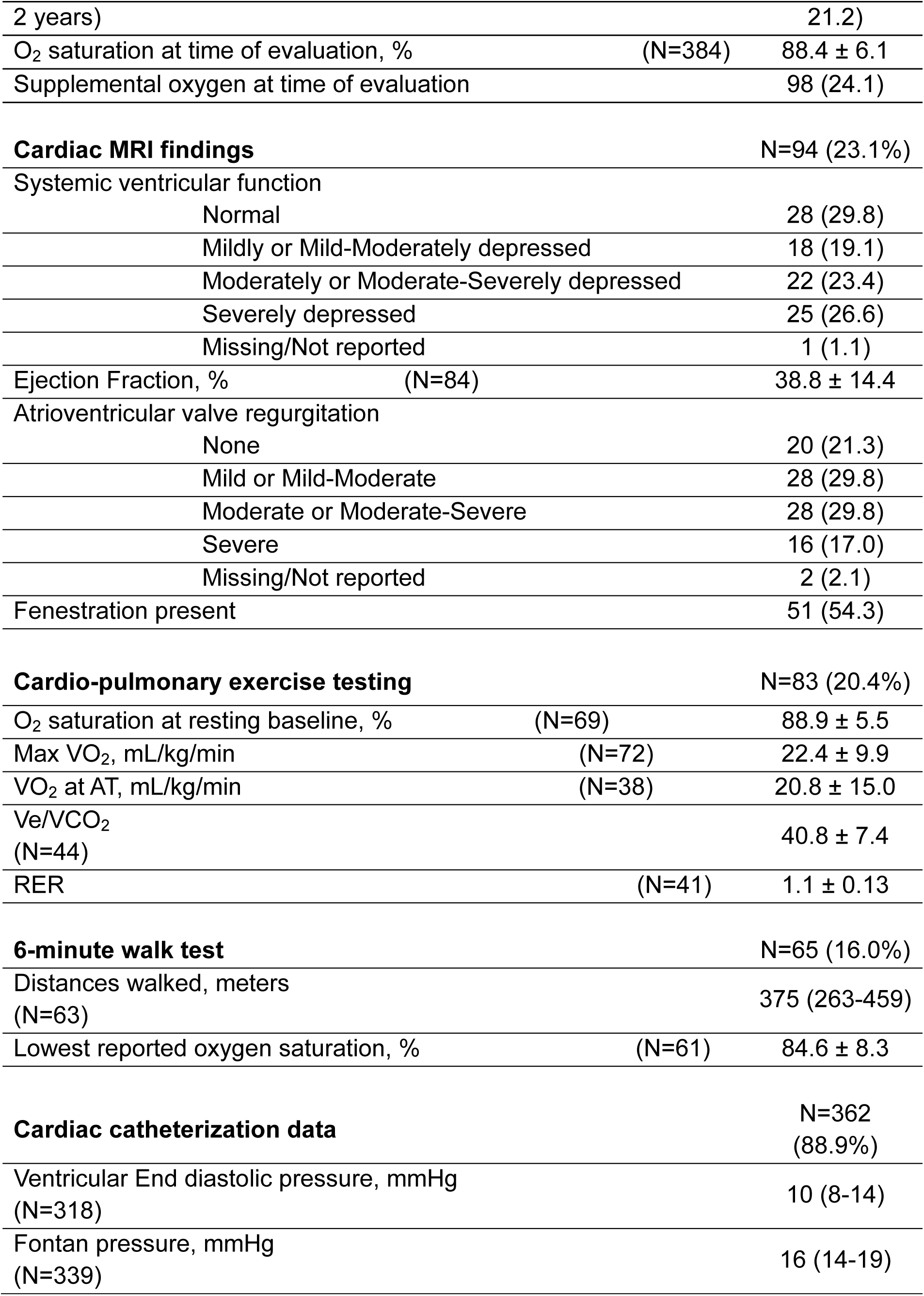
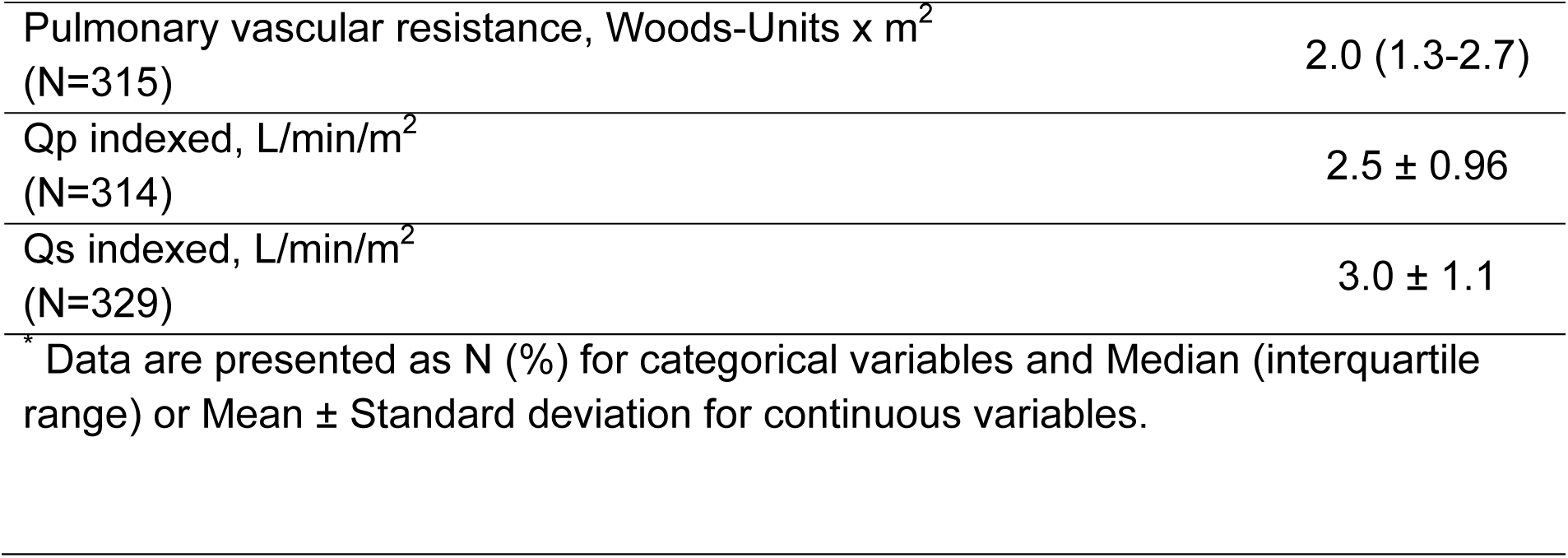
Transplant listing and Pre-transplant testing characteristics (N=409)

|  |  |  |
| --- | --- | --- |
| Age at Listing, years | (N=407) | 13.0 (6.5-16.6) |
| Time from Fontan to listing, years | (N=399) | 9.2 (2.8-13.1) |
| Status at listing |  |  |
| 1A |  | 153 (37.4) |
| 1B |  | 130 (31.8) |
| 2 |  | 77 (18.8) |
| Adult listing |  | 33 (8.1) |
| Canadian listing |  | 16 (3.9) |
| NYHA class |  |  |
| I |  | 28 (6.8) |
| II |  | 119 (29.1) |
| III |  | 143 (35.0) |
| IV |  | 90 (22.0) |
| Unknown |  | 29 (7.1) |
| Insurance type at time of evaluation |  |  |
| None |  | 2 (0.5) |
| Public |  | 211 (51.6) |
| Private |  | 170 (41.6) |
| Unknown |  | 26 (6.4) |
| Patient enrolled in school or employed at time of evaluation |  | 307 (75.1) |
| Single parent home at time of evaluation |  | 91 (22.2) |
| Parent/caregiver has a mental health history of any of the following (anxiety, mood, trauma, or substance use disorder) |  | 74 (18.1) |
| History of child protective service involvement |  | 32 (7.8) |
| Number of hospitalizations in the year prior to transplant listing |  | 1 (0-3) |
| 0 |  | 101 (24.7) |
| 1 |  | 105 (25.7) |
| 2-3 |  | 103 (25.2) |
| 4-5 |  | 43 (10.5) |
| 6+ |  | 11 (2.7) |
| Unknown |  | 46 (11.2) |
| Weight-for-age z-score at time of evaluation (N=360 for 2 < age < 20 years) |  | -1.0 ± 1.6 |
| Height-for-age z-score at time of evaluation (N=359 for 2 < age < 20 years) |  | -1.5 ± 1.5 |
| BMI at time of evaluation, kg/m<sup>2</sup> (N=397 for age > |  | 18.3 (16.2- |
| 2 years) |  | 21.2) |
| O<sub>2</sub> saturation at time of evaluation, % | (N=384) | 88.4 ± 6.1 |
| Supplemental oxygen at time of evaluation |  | 98 (24.1) |
| **Cardiac MRI findings** |  | N=94 (23.1%) |
| Systemic ventricular function |  |  |
| Normal |  | 28 (29.8) |
| Mildly or Mild-Moderately depressed |  | 18 (19.1) |
| Moderately or Moderate-Severely depressed |  | 22 (23.4) |
| Severely depressed |  | 25 (26.6) |
| Missing/Not reported |  | 1 (1.1) |
| Ejection Fraction, % | (N=84) | 38.8 ± 14.4 |
| Atrioventricular valve regurgitation |  |  |
| None |  | 20 (21.3) |
| Mild or Mild-Moderate |  | 28 (29.8) |
| Moderate or Moderate-Severe |  | 28 (29.8) |
| Severe |  | 16 (17.0) |
| Missing/Not reported |  | 2 (2.1) |
| Fenestration present |  | 51 (54.3) |
| **Cardio-pulmonary exercise testing** |  | N=83 (20.4%) |
| O<sub>2</sub> saturation at resting baseline, % | (N=69) | 88.9 ± 5.5 |
| Max VO<sub>2</sub>, mL/kg/min | (N=72) | 22.4 ± 9.9 |
| VO<sub>2</sub> at AT, mL/kg/min | (N=38) | 20.8 ± 15.0 |
| Ve/VCO<sub>2</sub> |  | 40.8 ± 7.4 |
| (N=44) |  |  |
| RER | (N=41) | 1.1 ± 0.13 |
| **6-minute walk test** |  | N=65 (16.0%) |
| Distances walked, meters |  | 375 (263-459) |
| (N=63) |  |  |
| Lowest reported oxygen saturation, % | (N=61) | 84.6 ± 8.3 |
| **Cardiac catheterization data** |  | N=362<br>(88.9%) |
| Ventricular End diastolic pressure, mmHg |  | 10 (8-14) |
| (N=318) |  |  |
| Fontan pressure, mmHg |  | 16 (14-19) |
| (N=339) |  |  |

|  |  |
| --- | --- |
| Pulmonary vascular resistance, Woods-Units x m<sup>2</sup><br>(N=315) | 2.0 (1.3-2.7) |
| Qp indexed, L/min/m<sup>2</sup><br>(N=314) | 2.5 ± 0.96 |
| Qs indexed, L/min/m<sup>2</sup><br>(N=329) | 3.0 ± 1.1 |

\* Data are presented as N (%) for categorical variables and Median (interquartile range) or Mean ± Standard deviation for continuous variables.

FCF-related morbidity characteristics of the cohort are shown in Table 3. The variation in FCF presentation within the cohort was broad. The median number of unique FCF- related morbidities present was 5 (IQR of 3-8 and a range of 0-15 morbidities), demonstrating significant heterogeneity in FCF presentation and triggers for heart transplant evaluation and waitlisting.

**Table 3.**
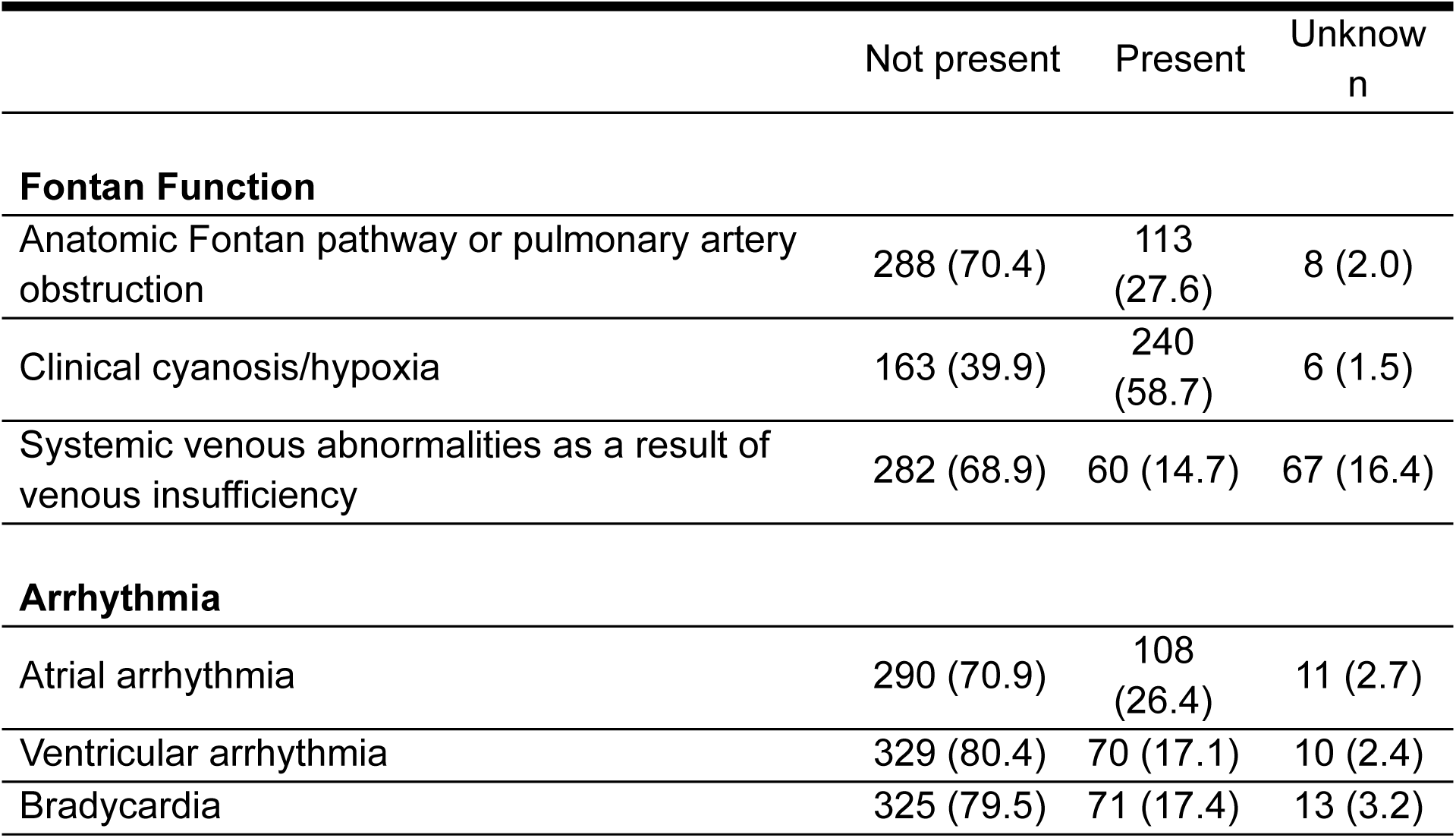
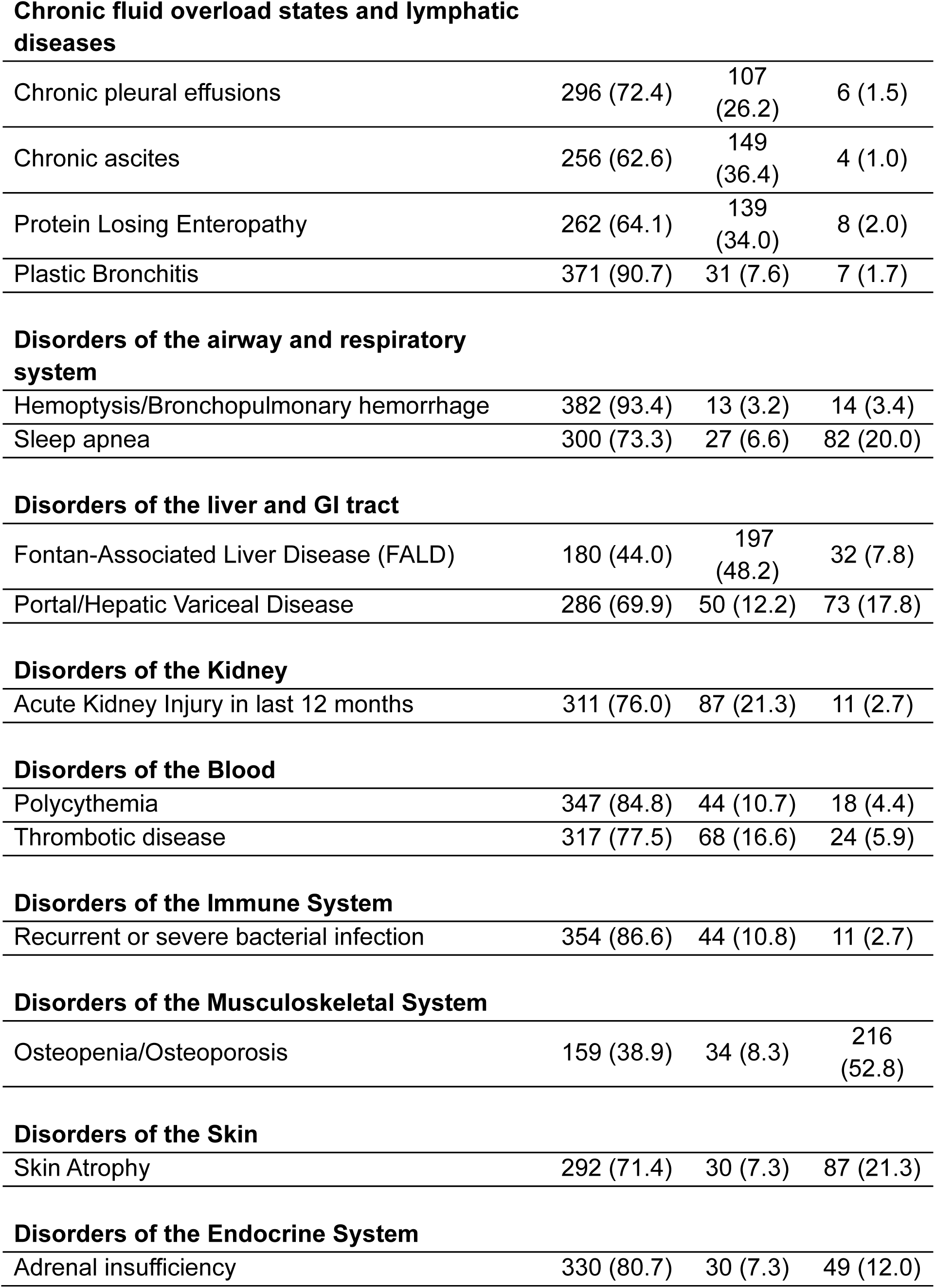
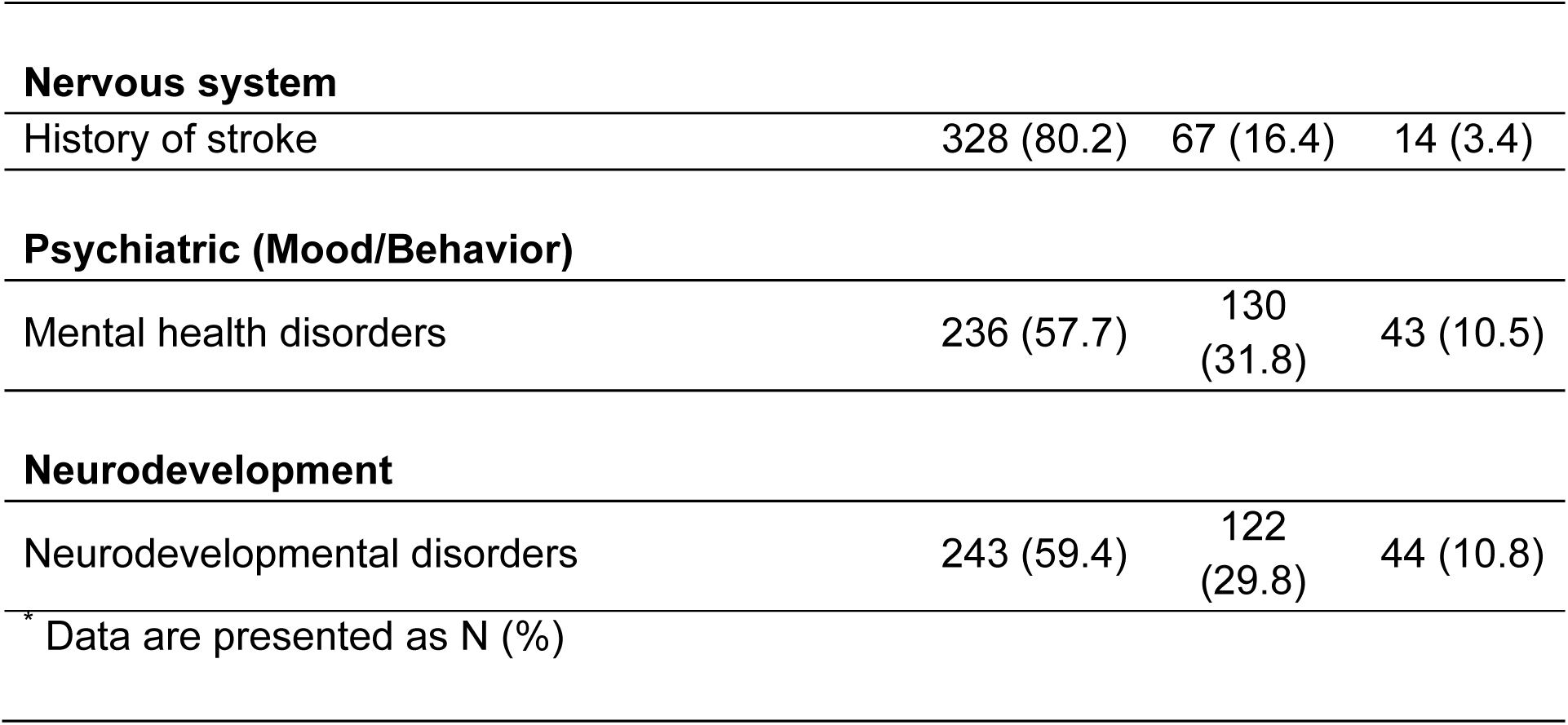
FCF-specific morbidities.

| **Table 3.** FCF-specific morbidities | Not present | Present | Unknown |
| --- | --- | --- | --- |
| **Fontan Function** |  |  |  |
| Anatomic Fontan pathway or pulmonary artery obstruction | 288 (70.4) | 113 (27.6) | 8 (2.0) |
| Clinical cyanosis/hypoxia | 163 (39.9) | 240 (58.7) | 6 (1.5) |
| Systemic venous abnormalities as a result of venous insufficiency | 282 (68.9) | 60 (14.7) | 67 (16.4) |
| **Arrhythmia** |  |  |  |
| Atrial arrhythmia | 290 (70.9) | 108 (26.4) | 11 (2.7) |
| Ventricular arrhythmia | 329 (80.4) | 70 (17.1) | 10 (2.4) |
| Bradycardia | 325 (79.5) | 71 (17.4) | 13 (3.2) |
| **Chronic fluid overload states and lymphatic diseases** |  |  |  |
| Chronic pleural effusions | 296 (72.4) | 107<br>(26.2) | 6 (1.5) |
| Chronic ascites | 256 (62.6) | 149<br>(36.4) | 4 (1.0) |
| Protein Losing Enteropathy | 262 (64.1) | 139<br>(34.0) | 8 (2.0) |
| Plastic Bronchitis | 371 (90.7) | 31 (7.6) | 7 (1.7) |
| **Disorders of the airway and respiratory system** |  |  |  |
| Hemoptysis/Bronchopulmonary hemorrhage | 382 (93.4) | 13 (3.2) | 14 (3.4) |
| Sleep apnea | 300 (73.3) | 27 (6.6) | 82 (20.0) |
| **Disorders of the liver and GI tract** |  |  |  |
| Fontan-Associated Liver Disease (FALD) | 180 (44.0) | 197<br>(48.2) | 32 (7.8) |
| Portal/Hepatic Variceal Disease | 286 (69.9) | 50 (12.2) | 73 (17.8) |
| **Disorders of the Kidney** |  |  |  |
| Acute Kidney Injury in last 12 months | 311 (76.0) | 87 (21.3) | 11 (2.7) |
| **Disorders of the Blood** |  |  |  |
| Polycythemia | 347 (84.8) | 44 (10.7) | 18 (4.4) |
| Thrombotic disease | 317 (77.5) | 68 (16.6) | 24 (5.9) |
| **Disorders of the Immune System** |  |  |  |
| Recurrent or severe bacterial infection | 354 (86.6) | 44 (10.8) | 11 (2.7) |
| **Disorders of the Musculoskeletal System** |  |  |  |
| Osteopenia/Osteoporosis | 159 (38.9) | 34 (8.3) | 216<br>(52.8) |
| **Disorders of the Skin** |  |  |  |
| Skin Atrophy | 292 (71.4) | 30 (7.3) | 87 (21.3) |
| **Disorders of the Endocrine System** |  |  |  |
| Adrenal insufficiency | 330 (80.7) | 30 (7.3) | 49 (12.0) |
| **Nervous system** |  |  |  |
| History of stroke | 328 (80.2) | 67 (16.4) | 14 (3.4) |
| **Psychiatric (Mood/Behavior)** |  |  |  |
| Mental health disorders | 236 (57.7) | 130 (31.8) | 43 (10.5) |
| **Neurodevelopment** |  |  |  |
| Neurodevelopmental disorders | 243 (59.4) | 122 (29.8) | 44 (10.8) |
| * Data are presented as N (%) |  |  |  |

Clinical status changed in a significant proportion of patients during the waitlisting period. Vasoactive infusions were initiated in 247 (60.4%) of patients prior to heart transplant and continued until the time of heart transplant or death in 82.6% of those patients. Mechanical ventilation was necessary in 57 (13.9%) patients after waitlisting and continued until transplant or death in 45.6% of ventilated patients. A new ventricular assist device was placed after waitlisting in 45 (11.0%) patients and remained in place in 91.1% of patients until transplant or death. Extracorporeal membrane oxygenation was used in 20 (4.9%) patients and continued until death or transplant in 50.0% of those patients.

Twenty-four (5.9%) patients died while waiting and 20 (4.9%) patients were removed from the waitlist. Of those removed from the waitlist,4 were removed because they were deemed to have too much morbidity/illness to remain a transplant candidate.

Additionally, 8 patients’ status improved to no longer needing transplant at the time, and 8 patients’ families requested to be removed from the waitlist or otherwise were unable to continue with transplant listing for reasons other than clinical status (these patients were censored from further analysis).

Figure 2 demonstrates the overall survival of waitlist patients. Notably, 24 (5.9%) patients remained on the waitlist for more than 1-year at the time of data collection. The following factors were associated with waitlist death in univariate analysis: higher grade (2+ or greater) of AP collaterals (28.6% vs. 15.3%, p=0.01), higher number of hospitalizations in the year preceding listing (hospitalization at the time of listing counted as 1 hospitalization; median 2 vs. 1, p=0.048), younger age at transplant evaluation (median 5.2 years vs. 13.1 years, p=0.003), sleep apnea (16.7% vs. 4.7%, p=0.04), higher NYHA class (class IV; 41.7% vs. 23.2%, p=0.01), non-enrollment in school or non-employment (45.8% vs. 17.9%, p=0.002), and single parent home at time of evaluation (37.5% vs. 20.5%, p=0.04).

**Figure 2.**
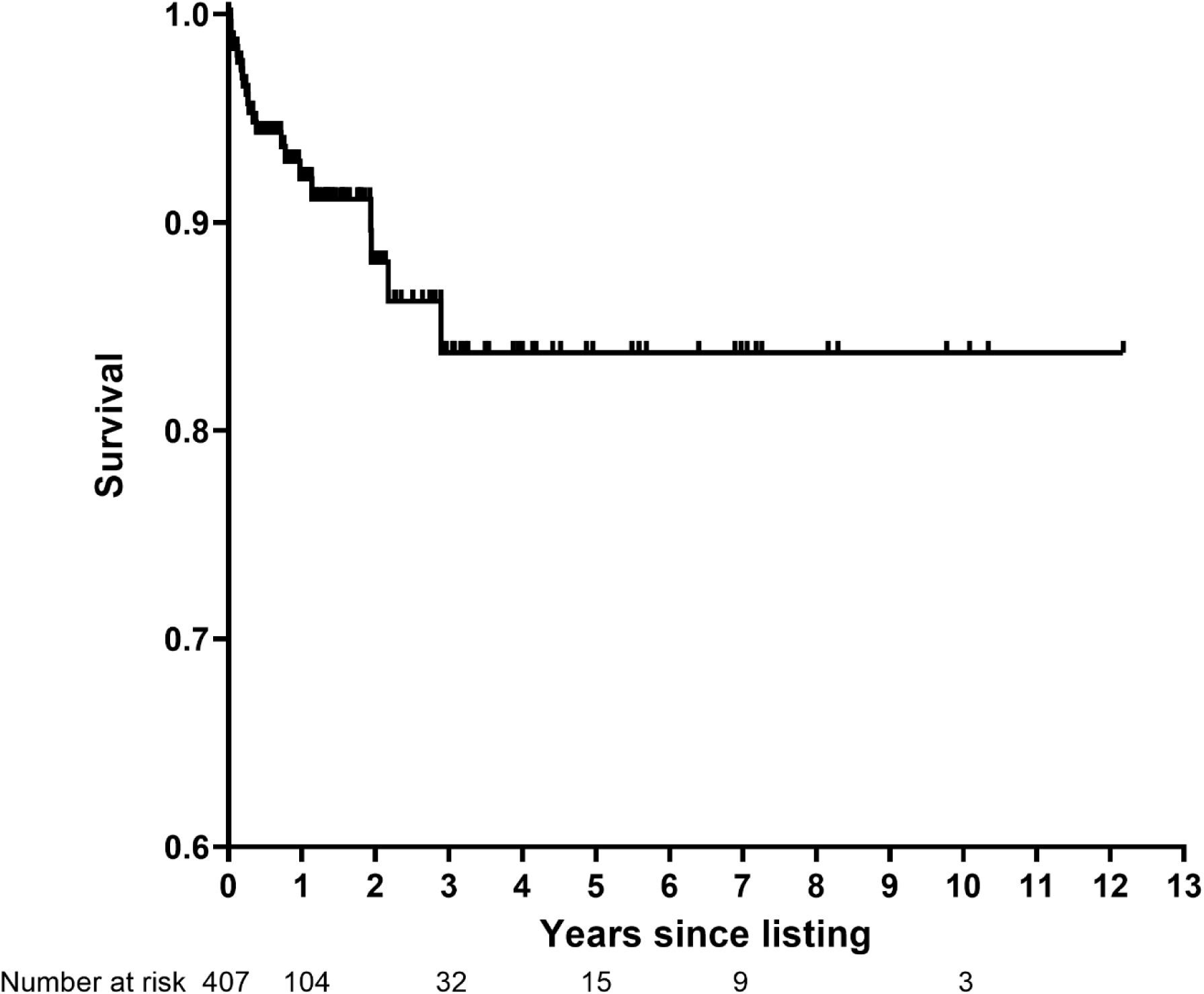
Waitlist survival censored at time of transplant or removal from the transplant waitlist for reasons other than being too ill for transplant candidacy. * Two patients with unknown date of listing were not included.

### Post-heart transplant survival and transplant morbidities

In the 341 (83.4%) patients undergoing heart transplant, the median post-transplant follow-up time was 4.2 years (IQR 1.9-7.2). Dual organ transplant occurred in 3.8% (N=13) of the cohort including 12 heart-liver transplants and 1 heart-kidney transplant; due to the low number of patients undergoing dual-organ transplant they were included in the overall cohort and not analyzed separately. A total of 27 (7.9%) patients died in the first-year post-transplant. An additional 5 individuals (1.7%) died between 1 and 3- years post-transplant. Survival in the first-year post-transplant is demonstrated in Figure 3. Those who died in the first post-transplant year were more likely to have a diagnosis of hypoplastic left heart syndrome as the type of single ventricle CHD (81.5% vs. 61.9%, p=0.04), normal systolic function on echocardiogram (55.6% vs. 33.1%, p=0.03), a patent fenestration (74.1% vs. 42.9%, p=0.002), a lower oxygen saturation (85.0 ± 5.7 vs. 88.9 ± 6.3, p=0.003), a supplemental oxygen requirement (44.4% vs. 24.4%, p=0.02), a higher class II PRA (median 3% vs. 0%, p=0.03), Fontan pathway or pulmonary artery obstruction (44.4% vs. 23.5%, p=0.02), clinical cyanosis defined as a resting arterial oxygen saturation < 90% (88.9% vs. 56.4%, p=0.001), higher grade portal variceal disease (25.9% vs. 8.0%, p=0.01), polycythemia (25.9% vs. 9.7%, p=0.02), and a mental health condition requiring treatment (51.9% vs. 28.0%, p=0.04).

**Figure 3.**
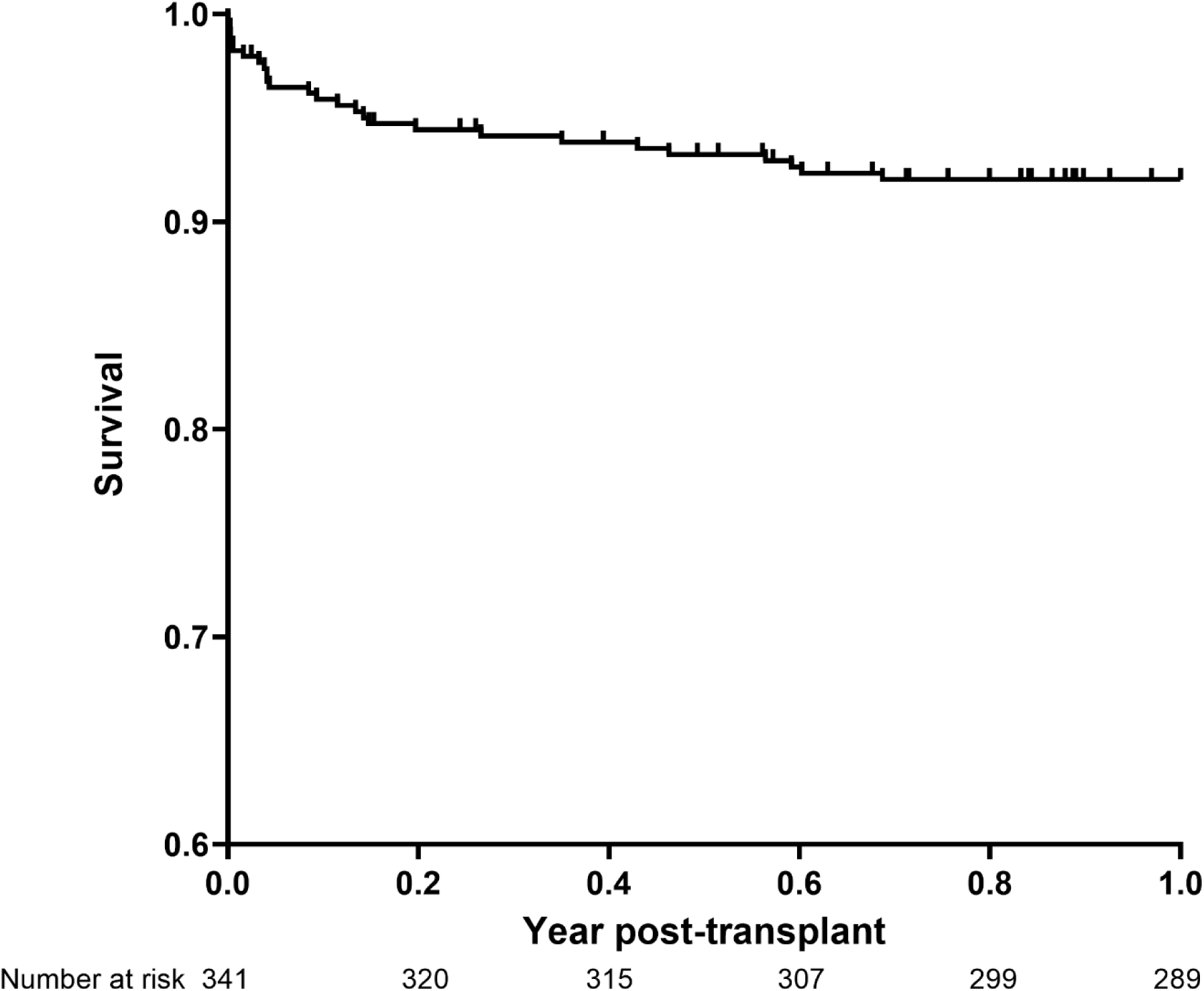
Survival to 1-year post-transplant conditional on survival to transplant.

In the first-year post-transplant, 38.9% of the cohort had allograft rejection requiring an adjustment in immunosuppressive therapy; 27.5% had cellular-mediated rejection and 22.8% had antibody-mediated rejection (non-exclusive). Neither type of rejection, nor rejection overall, was associated with 1-year mortality (25.9% in deaths vs 40.1% in survivors, p=0.14). Bacterial infection requiring parenteral antibiotics occurred in 22.2% of the cohort in the first-year post-transplant; 37.0% of the patients that died had an infection compared to 20.8% of survivors (p=0.06). Fungal infections were rarer but occurred in 5.7% of the cohort including 18.5% of those who died vs 4.5% of survivors (p=0.01). Post-transplant bacterial infections were more likely to have occurred in those who also had recurrent or severe bacterial infections prior to transplant (55.3% vs. 19.4%; p<0.0001).

### Overall survival from time of waitlisting to 1-year post-heart transplant

For the primary outcome of survival from waitlist to 1-year post-transplant, 316 patients met an outcome of either 1-year survival (n=289) or mortality (n=27). At the time of data analysis, 24 were still on the waitlist at last follow-up, 25 were alive but had less than 1- year post-HT follow-up, and 20 were censored at the time of removal from the waitlist for reasons other than clinical deterioration. Among the 15.0% (n=27) of individuals who died, after seeindependent risk factors for mortality included >1 hospitalization in the year prior to waitlisting (aOR 2.0, p=0.05) and clinical cyanosis (aOR 5.0, p=0.002) (Figure 4). Overall survival (including beyond 1-year post-transplant) since waitlisting for the cohort is demonstrated in Figure 5.

**Figure 4.**
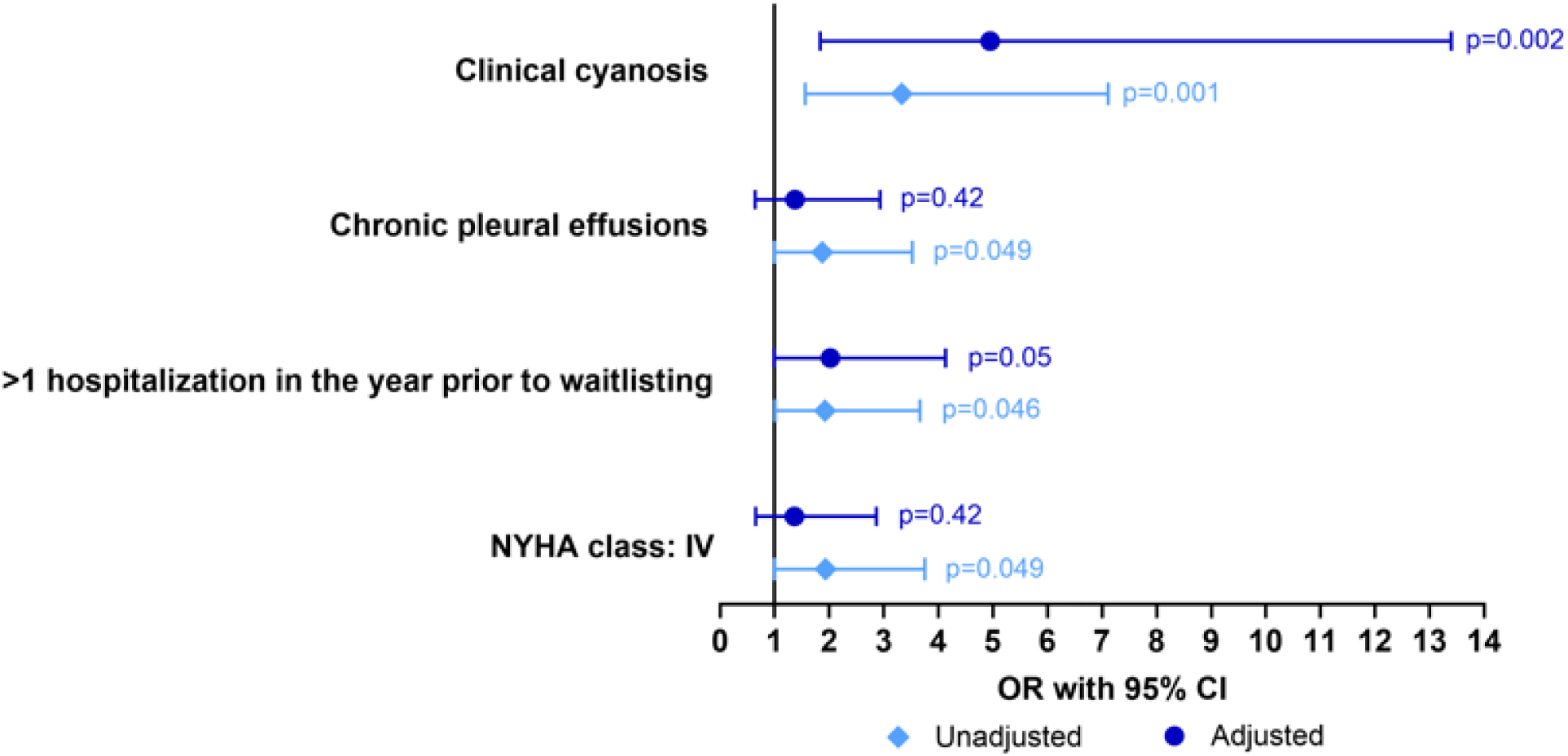
Forest plot comparison of risk factors in the final multivariable model for non-survival from transplant to 1-year post-operative with factors affecting survival from waitlisting until 1-year post-transplant.

**Figure 5.**
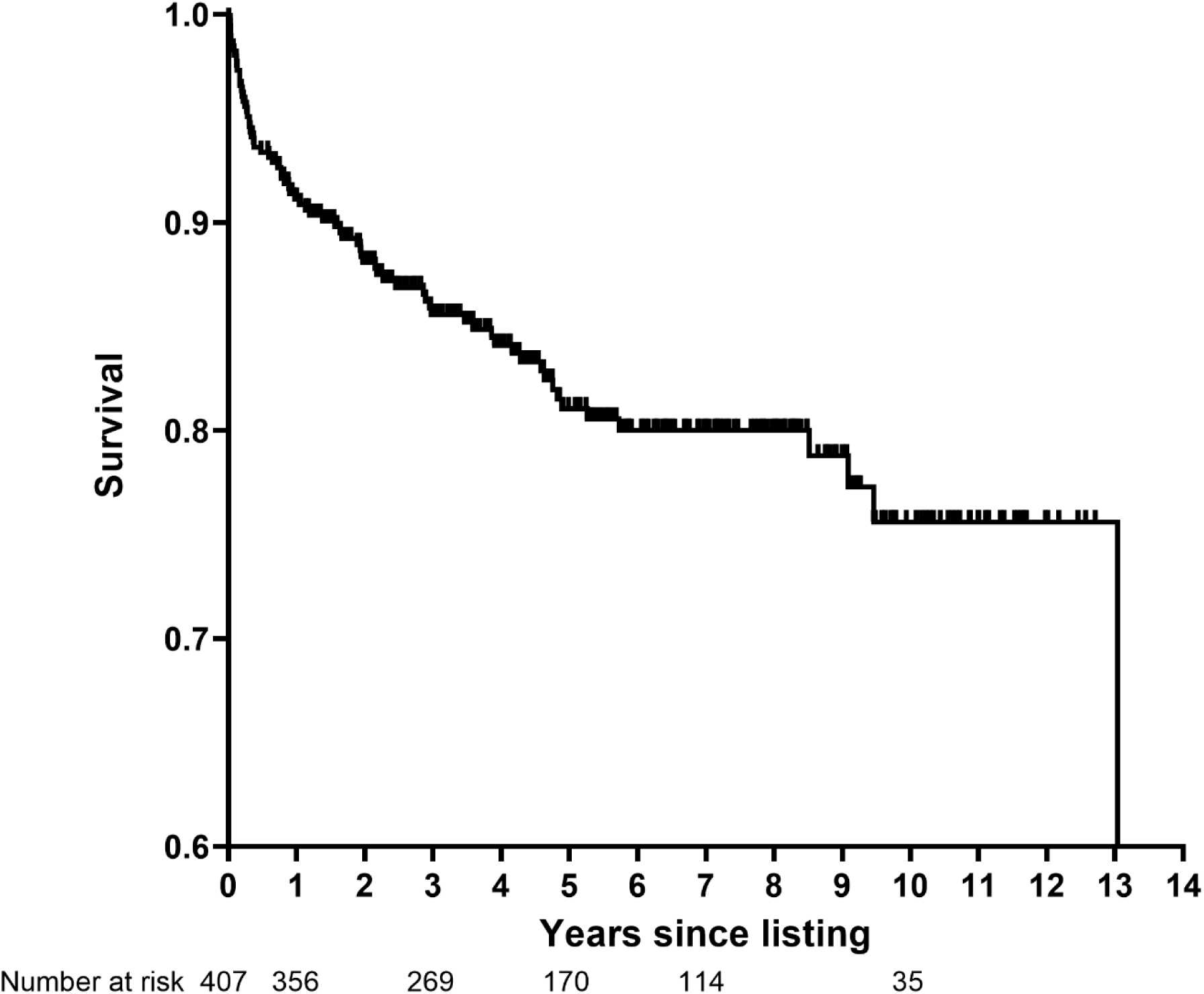
Overall survival since waitlisting. * Two patients with unknown date of listing were not included.

## Discussion

This study provides the largest, most comprehensive FCF-specific report of waitlist and post-heart transplant outcomes in individuals with Fontan physiology. Survival of Fontan patients selected for transplant both on the waitlist and early post-transplant among expert congenital heart centers was notably higher in this recent era cohort than previously seen which suggests significant gains in outcomes and improved understanding of this population as a field. Compared to other multicenter reports of transplant listing, waitlist mortality has improved to 5.9 % from previously reported 12- 26%, and 1-year post transplant survival has improved to 92.1% from previously reported 76%-89% [3, 4, 8, 9] . However, there remains significant mortality from the time of waitlist to 1-year post-transplant that may still be improved upon, and unique risk factors were determined via this study to help guide referral and treatment.

Risk factors for death on the waitlist and death post-transplant were remarkably different. Patients who died while waiting were more likely to have worsened classical symptoms of heart failure as evidenced by higher NYHA class. This may be exacerbated by worsened AP collateralization, also a risk factor, that can complicate systemic cardiac output and oxygen delivery. They were also more likely to admitted to the hospital repeatedly in the year prior to transplant, again likely driven by acute symptoms. In contrast, those surviving the waiting period but dying early post-transplant were more likely to have normal ventricular function by echo and less severe typical heart failure symptoms but instead had other clinical findings that reflect the more indolent and slowly progressive failure of the Fontan pathway and dysfunction from chronically elevated central venous pressures. They were more likely to have Fontan pathway obstruction, increased cyanosis, and portal varices. These findings suggest chronic, lifetime changes that are not immediately reversed by transplantation impact outcome even after restoration of biventricular physiology which is consistent with other phenotypic characterizations of Fontan patients undergoing advanced therapies [10]. When waitlist and post-transplant risk factors are viewed together, they present a complex picture for evaluation and monitoring in the Fontan patient. However, one can conclude from this study, that acute new symptoms as well as progression of chronic effects of Fontan circulation must both be continually assessed and when present, referral to advanced heart failure teams should be considered.

While cyanosis has been invoked as an association with worsened outcomes in some non-transplant-oriented studies [11, 12], its independent association with overall mortality after listing for transplant in this study is novel and pronounced. While the only other independent risk factor for mortality, multiple hospitalizations, may be a catch-all for many severe forms of rarer FCF presentation, cyanosis is a single, easily measurable clinical finding. It is not immediately clear why cyanosis is associated with impaired survival. Cyanosis may be a marker of more deranged Fontan physiology. It may be that chronic cyanosis causes systemic changes that lead to multiorgan disease or predispose to perioperative morbidity such as bleeding. Potentially cyanosis also leads to lifetime exercise impairment and deconditioning that affects ability to survive the physical challenges of transplantation. Further, ongoing post-transplant cyanosis may have direct effects on the graft. The causes that lead to cyanosis at time of waitlisting being an independent risk factor warrants further study.

This study also serves as a detailed characterization FCF itself in a large cohort. In short, FCF and its clinical presentation are remarkably heterogeneous. The median number of unique FCF factors in the cohort was 5 with an IQR of 3 to 8 factors. This illustrates the challenges clinicians face in monitoring patients with Fontan physiology.

These individuals develop multiple clinical abnormalities all of which may change or worsen over time. How FCF morbidities should be tracked and what thresholds or combinations of factors should trigger transplant require additional study. In the future, we plan to study this cohort in detail to assess combinations of FCF morbidities and overall disease burden to look for presentation patterns that may help to guide surveillance testing, referral, and evaluation for advanced heart failure therapies.

Finally, the impact of both psychosocial factors and social drivers of health must be acknowledged. Both single parent household and non-enrollment in school or employment were risk factors for waitlist mortality but neither were associated with post-transplant mortality. It is unclear whether this finding is related to the individual’s degree of heart failure or not. We fully recognize that these chart-based variables are not comprehensive measures of social drivers of health, however, they may serve as an indicator of possible inequities in access to care or historical marginalization. Similarly to others who have discerned race-based and socio-economic disparities in waitlist outcomes [13–15], these findings underscore the importance of prioritizing early identification of and referral for heart failure therapy particularly among patients from disadvantaged communities.

Notably, the presence of mental health condition(s) requiring treatment as indicated in the medical chart was found to negatively impact post-transplant survival. In fact, the risk of post-transplant mortality was almost doubled in those with mental health conditions requiring treatment. Given the higher rates of mental health diagnoses among the Fontan-palliated heart disease population [16–18] coupled with established associations between mental health comorbidity and post-transplant treatment adherence [19], regular surveillance and early referral for mental health care needs are as important as physical FCF morbidity monitoring. While several FCF morbidities are less modifiable to intervention, mental health represents a risk factor that can be prevented or mitigated to improve post-transplant outcomes. Prevention-focused and integrated psychosocial care is critical for the patient with Fontan-palliation and should be prioritized at institutional and program levels to improve outcomes in this higher risk group.

This study has several limitations. First, this is a retrospective cohort study and while care was taken to collect data that could be obtained from chart review, many FCF morbidities and other information may not be completely available from chart review. Relatedly, some chart-based variables, such as single parent household and presence of a mental health condition, do not fully capture the complexity of these constructs, such as additional social drivers of health or type of mental health condition impacting outcomes. Second, despite 20 centers and more than 400 patients, the overall mortality was relatively low which compromises power to detect risk factors that may be still important. Further, although 20 centers were included, they were chosen specifically as centers with significant Fontan transplant volume and experience and the findings may not be generalizable. Additionally, some findings, such as PRA, echocardiographic, and angiographic findings may have site specific measures without real normalization of values. There is also no information provided by this study regarding initial patient selection for waitlisting; data regarding patients who were declined for transplant is not present and our understanding of this group remains very limited. Finally, other markers of overall physical and mental health, such as physical frailty or psychological resilience are commonly gauged as part of patient evaluation for advanced therapies. Those indicators representing an individual’s overall response to illness may be as or more predictive than any individual morbidity.

These limitations inform the next phase in this critically important field of research given the growing number of Fontan transplants. The severity of individual FCF morbidities requires further study. Further, some patient factors, such as collateral vessels, may be modifiable, but whether modification affects outcome is unknown. Specific evaluation of frailty and resilience, both of which may be modified to improve outcomes in other disease states, is of significant interest.

## Conclusion

Though improved compared to previous reports, Fontan-palliated patients selected for transplant continue to have high mortality from waitlisting through transplant. Among FCF specific morbidities, cyanosis is associated with worsened survival. The presentation of FCF is very heterogeneous and warrants continued study to determine specific Fontan patient surveillance and referral.

## Data Availability

Datasets are unable to be published due to terms of existing data use agreements between centers and identifiable patient information.

## Acknowledgements

Data collected by the Pediatric Cardiac Critical Care Consortium and Western Canadian Single Ventricle Registry were used for this manuscript with permission from each organization. The authors would also like to acknowledge the research staff at each center that assisted in this effort.

## REFERENCES

1. Allen, K.Y., et al., Effect of Fontan-Associated Morbidities on Survival With Intact Fontan Circulation. Am J Cardiol, 2017. 119(11): p. 1866–1871.

2. Goldberg, D.J., et al., The failing Fontan: etiology, diagnosis and management. Expert Rev Cardiovasc Ther, 2011. 9(6): p. 785–93.

3. Bernstein, D., et al., Outcome of listing for cardiac transplantation for failed Fontan: a multi-institutional study. Circulation, 2006. 114(4): p. 273–80.

4. Simpson, K.E., et al., Fontan Patient Survival After Pediatric Heart Transplantation Has Improved in the Current Era. Ann Thorac Surg, 2017. 103(4): p. 1315–1320.

5. Chen, S., et al., Outcomes after initial heart failure consultation in Fontan patients. Cardiol Young, 2024. 34(5): p. 989–996.

6. Lewis, M.J., et al., Clinical Outcomes of Adult Fontan-Associated Liver Disease and Combined Heart-Liver Transplantation. J Am Coll Cardiol, 2023. 81(22): p. 2149–2160.

7. Schumacher, K.R., et al., Achieving Consensus: Severity-Graded Definitions of Fontan-Associated Complications to Characterize Fontan Circulatory Failure. J Card Fail, 2024.

8. Kovach, J.R., et al., Comparison of risk factors and outcomes for pediatric patients listed for heart transplantation after bidirectional Glenn and after Fontan: an analysis from the Pediatric Heart Transplant Study. J Heart Lung Transplant, 2012. 31(2): p. 133–9.

9. Kulshrestha, K., et al., The majority of pediatric Fontan patients have excellent post-transplant survival. J Thorac Cardiovasc Surg, 2024. 167(6): p. 2193–2203.

10. Dykes, J.C., et al., Clinical and hemodynamic characteristics of the pediatric failing Fontan. J Heart Lung Transplant, 2021. 40(12): p. 1529–1539.

11. Schafstedde, M., et al., Persisting and reoccurring cyanosis after Fontan operation is associated with increased late mortality. Eur J Cardiothorac Surg, 2021. 61(1): p. 54–61.

12. Egbe, A.C., et al., Predictors of procedural complications in adult Fontan patients undergoing non-cardiac procedures. Heart, 2017. 103(22): p. 1813–1820.

13. Bansal, N., et al., Impact of race and health coverage on listing and waitlist mortality in pediatric cardiac transplantation. J Heart Lung Transplant, 2023. 42(6): p. 754–764.

14. Amdani, S., A. Tang, and J.D. Schold, Children from socioeconomically disadvantaged communities present in more advanced heart failure at the time of transplant listing. J Heart Lung Transplant, 2023. 42(2): p. 150–155.

15. Singh, T.P., et al., Association of race and socioeconomic position with outcomes in pediatric heart transplant recipients. Am J Transplant, 2010. 10(9): p. 2116–23.

16. DeMaso, D.R., et al., Psychiatric Disorders in Adolescents With Single Ventricle Congenital Heart Disease. Pediatrics, 2017. 139(3).

17. McCormick, A.D., et al., Psychological functioning in paediatric patients with single ventricle heart disease: a systematic review. Cardiol Young, 2022. 32(2): p. 173–184.

18. Gonzalez, V.J., et al., Mental Health Disorders in Children With Congenital Heart Disease. Pediatrics, 2021. 147(2).

19. Killian, M.O., et al., Psychosocial predictors of medication non-adherence in pediatric organ transplantation: A systematic review. Pediatr Transplant, 2018. 22(4): p. e13188.

